# Hidden heterogeneity and its influence on dengue vaccination impact

**DOI:** 10.1101/19006783

**Authors:** Magdalene K. Walters, T. Alex Perkins

## Abstract

The CYD-TDV vaccine was recently developed to combat dengue, a mosquito-borne viral disease that afflicts millions of people each year throughout the tropical and subtropical world. Its rollout has been complicated by recent findings that vaccinees with no prior exposure to dengue virus (DENV) experience an elevated risk of severe disease in response to their first DENV infection subsequent to vaccination. As a result of these findings, guidelines for use of CYD-TDV now require serological screening prior to vaccination to establish that an individual does not fall into this high-risk category. These complications mean that the public health impact of CYD-TDV vaccination is expected to be higher in areas with higher transmission. One important practical difficulty with tailoring vaccination policy to local transmission contexts is that DENV transmission is spatially heterogeneous, even at the scale of neighborhoods or blocks within a city. This raises the question of whether models based on data that average over spatial heterogeneity in transmission could fail to capture important aspects of CYD-TDV impact in spatially heterogeneous populations. We explored this question with a deterministic model of DENV transmission and CYD-TDV vaccination in a population comprised of two communities with differing transmission intensities. Compared to the full model, a version of the model based on the average of the two communities failed to capture benefits of targeting the intervention to the high-transmission community, which resulted in greater impact in both communities than we observed under even coverage. In addition, the model based on the average of the two communities substantially overestimated impact among vaccinated individuals in the low-transmission community. In the event that the specificity of serological screening is not high, this result suggests that models that ignore spatial heterogeneity could overlook the potential for harm to this segment of the population.

## INTRODUCTION

Dengue is a viral, mosquito-borne disease caused by any of four dengue virus (DENV) serotypes that are transmitted among people by *Aedes aegypti* and *Ae. albopictus* mosquitoes (1). While any of the four DENV serotypes may cause disease, an individual’s second infection, regardless of DENV serotype, carries an enhanced risk of causing severe disease (2; 3). This can result in hemorrhagic fever, hospitalization, and, in some cases, death (1). Due to the widespread distribution of its mosquito vectors (4; 5) dengue has been estimated to pose a risk to a large fraction of the world’s population, both now and in the future (6).

Despite the compelling need for a vaccine against dengue (7), only one dengue vaccine (CYD-TDV, or Dengvaxia, by Sanofi Pasteur) has been licensed to date. The rollout of this vaccine has been controversial (8), given that it has been associated with an elevated risk of severe disease for individuals who experience their first natural DENV infection at some point after being vaccinated (9). As a result, the Strategic Advisory Group of Experts on Immunization (SAGE) recommends that CYD-TDV only be administered to individuals with known prior DENV exposure (10). It is generally accepted that pre-vaccination serological screening is required to determine whether individuals meet this criterion (8). As such, we refer to the combination of serological screening and–in the event of a positive screening result–vaccination as the “intervention.”

There are three considerations that affect the public health impact of CYD-TDV vaccination following positive serological screening, each of which depends on baseline seroprevalence in the population being vaccinated. First, it is essential that screening have high specificity to avoid harm to seronegative individuals who should not be vaccinated, particularly in areas with low baseline seroprevalence (11). Second, it is also important that screening have high sensitivity to ensure that seropositive individuals benefit from vaccination, particularly in areas with high baseline seroprevalence (12). Third, an assay for serological screening with a given sensitivity and specificity is expected to have greater impact in an area with higher baseline seroprevalence, due to the higher proportion of seropositive individuals and the greater benefits of herd immunity in these settings (13). For these reasons, baseline seroprevalence could influence decisions about which screening assay is most appropriate in a given setting, or whether vaccination with CYD-TDV is appropriate in the first place.

One major complication to the formulation of policies for CYD-TDV vaccination is the fact that baseline seroprevalence can vary considerably across space (14). Among districts within a country, estimates of seroprevalence among nine-year-olds varied from below 20% to above 80% across departments in Colombia and states in Mexico, due to subnational variability in environmental drivers of transmission (11). Even below the level of a city, baseline seroprevalence can vary across neighborhoods or blocks. DENV seroprevalence ranged 56-77% across neighborhoods of Rio de Janeiro, Brazil (15),74-91% across neighborhoods of Recife, Brazil (16), 67-90% across neighborhoods in Iquitos, Peru (17), and 21-100% between adjacent blocks in Maracay, Venezuela (18). Drivers of variability in DENV seroprevalence at this spatial scale can include a number of factors associated with socioeconomic differences, such as housing quality, free-standing water, open containers, and infrequent waste removal (19; 20; 21; 22).

In the event that the spatial resolution of DENV seroprevalence estimates to inform CYD-TDV policy is limited to larger administrative scales (11), there is a risk that such policies may not be well-suited to constituent areas where seroprevalence is above or below average. Under the assumption of uniform seroprevalence, CYD-TDV implementation could pose a greater risk of severe disease in areas where seroprevalence is below average. Conversely, assuming uniform seroprevalence could lead to underestimates of the benefits from vaccination in areas of higher than average seroprevalence. Another possibility is that individuals from low-transmission communities could benefit indirectly from vaccination in high-transmission communities (23). Yet another complication is that vaccination coverage could vary spatially. In urban areas of Brazil, vaccination coverage has been found to be both negatively (24) and positively (25) associated with socioeconomic status, due to differences in access to vaccination and willingness to vaccinate across socioeconomic strata.

To date, mathematical models of CYD-TDV impact have mostly assumed that transmission is spatially homogeneous within the population being modeled (26). This leaves an important gap in understanding the implications of uniform policies for CYD-TDV vaccination in areas comprised of communities with unrecognized spatial heterogeneity in baseline DENV seroprevalence. To explore these implications, we developed and analyzed a simple two-patch model of DENV transmission based on a system of ordinary differential equations. Transmission potential and baseline seroprevalence differed between these hypothetical communities, and routine vaccination with CYD-TDV following serological screening in nine-year-olds was simulated over a thirty-year horizon. We examined differences in the proportion of cases averted among vaccinated and unvaccinated individuals in both communities, including under differing assumptions about human mobility and uneven vaccination coverage.

## METHODS

### Model Description

#### Overview

Our model tracks the transmission dynamics of DENV in two communities indexed by *c*. These communities, *h* and *l*, are differentiated by their respective high and low transmission coefficients, *β*_*h*_ and *β*_*l*_. They are coupled by human mobility, with individuals in susceptible classes assumed to spend a fraction, *H*, of their time in the community in which they reside and the rest of their time, 1-*H*, in the other community. Our model also included demographic change, as we are interested in modeling transmission over several decades. This includes a year-specific birth rate, μ_*t*_, year- and age-specific death rate, δ_*t,a*_, and progression through age classes 0-80+, indexed by *a*.

Rather than track all four DENV serotypes explicitly (something very few DENV transmission models do, (27)), our model stratifies individuals by how many DENV serotypes, *s*, they have been infected by. Individuals are further classified with respect to their status as susceptible, infected, or recovered within each community and age class, resulting in state variables *S*_*c,a,s*_, *I*_*c,a,s*_, and *R*_*c,a,s*_. Individuals in different *s* strata experience different probabilities of symptomatic disease, p_*s*_. As assumed by other models examining CYD-TDV impacts on DENV transmission (28), we incremented *s* by one upon vaccination. Upon vaccination, individuals were moved to unique states for vaccine recipients that were stratified similarly: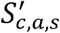, 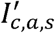 and 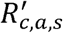.

Model parameters are summarized in Table 1, and a schematic representation of the order in which transitions among state variables occur is presented in Fig. 1. Code for numerically solving the system of ordinary differential equations (ODEs) associated with this model is available at https://github.com/mwalte10/intra-urban_dengue_vaccination_impact. This code made use of the ode45 function with default settings in the deSolve package (29) in the R programming language (30).

**Table 1.** Model parameters. The timescale of all rate parameters is per day.

| Parameter | Definition | Value | Source |
| --- | --- | --- | --- |
| $\mu_y$ | Birth rate in year $y$ | $2.8 \times 10^{-5}$ - $1.2 \times 10^{-4}$ | (31) |
| $\delta_{y,a}$ | Death rate in year $y$ at age $a$ | $4.6 \times 10^{-7}$ - $1.2 \times 10^{-3}$ | (31) |
| $H$ | Proportion of time spent in community of residence (travel, no travel) | 0.975, 1 | Assumed |
| $\beta_h$ | Transmission coefficient (travel, no travel) | 0.253, 0.245 | Calibrated |
| $\beta_l$ | Transmission coefficient (travel, no travel) | 0.127, 0.175 | Calibrated |
| $\bar{v}_c$ | Average intervention coverage across communities $h$ and $l$ | 0.5 | Assumed |
| $v_c$ | Intervention coverage in community $c$ | 0 - 1 | Varied |
| $sens$ | Sensitivity of pre-vaccination screening | 0.95 | (13) |
| $spec$ | Specificity of pre-vaccination screening | 0.85 | (13) |
| $\gamma$ | Rate of recovery from infection | 0.25 | (32) |
| $\sigma$ | Rate of loss of cross-immunity | 0.0023 | (26) |
| $\rho_s$ | Proportion of DENV infections that result in symptomatic disease ($s = 0, 1, 2, 3$) | 0.53, 1, 0.115, 0.115 | (26) |

**Figure 1.**
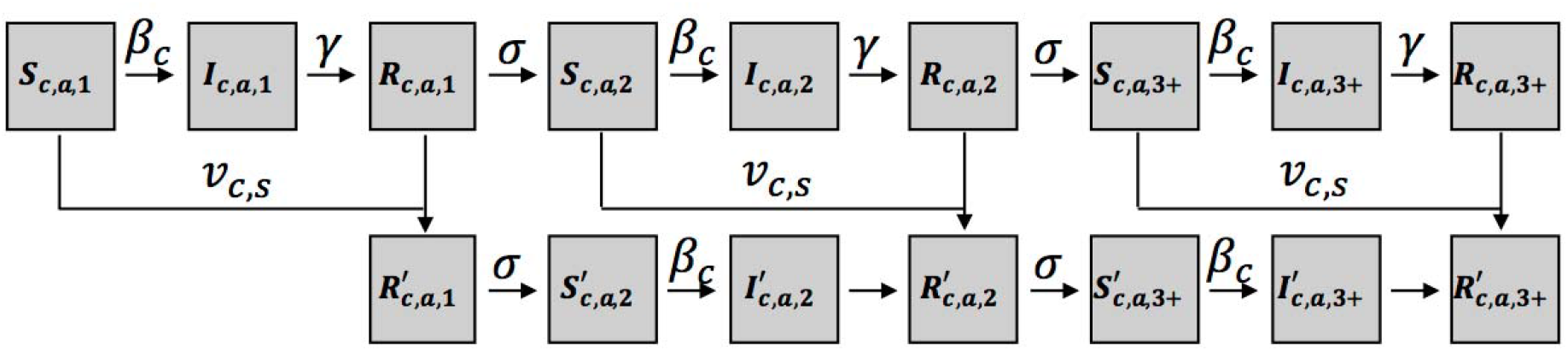
Schematic representation of transitions (arrows) among state variables (boxes). Demographic parameters and human mobility are excluded for simplicity.

#### Demography

Our model tracked ages in one-year increments from zero to 79, and individuals aged 80 or older were binned into a single group. Transitions into ages one to 80+ occurred at a daily rate of *G*_*a*_ = 365^-1^. Transitions out of ages zero to 79 occurred at rate 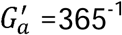, while individuals in the terminal age class of 80+ years remained there until death. Births occurred at year-specific, daily rate μ_*t*_ per capita, which was multiplied by total population *N*_*c*_(*t*) to obtain the overall rate at which newborns entered community *c* through the *S*_*c,0,1*_ compartment. Deaths occurred at year- and age-specific, daily rate δ_*t,a*_ for individuals in all states. The background death rate, δ_*t,a*_, was assumed to adequately capture the small proportion of DENV infections that resulted in death. Although our model was not designed with a specific population in mind, we used birth and death rates from Brazil (31) given that it is a large, dengue-endemic country that has been the subject of CYD-TDV modeling before (13; 26). To ground our analysis in contemporary demographic rates, we assumed that CYD-TDV vaccination began in 2020, meaning that the model burn-in period began in 1960 and that vaccination impacts were tracked until 2050. Over the course of this timeframe, birth rate decreased more than four-fold (Fig. 2A), and child mortality decreased while mortality among older ages increased (Fig. 2B).

**Figure 2.**
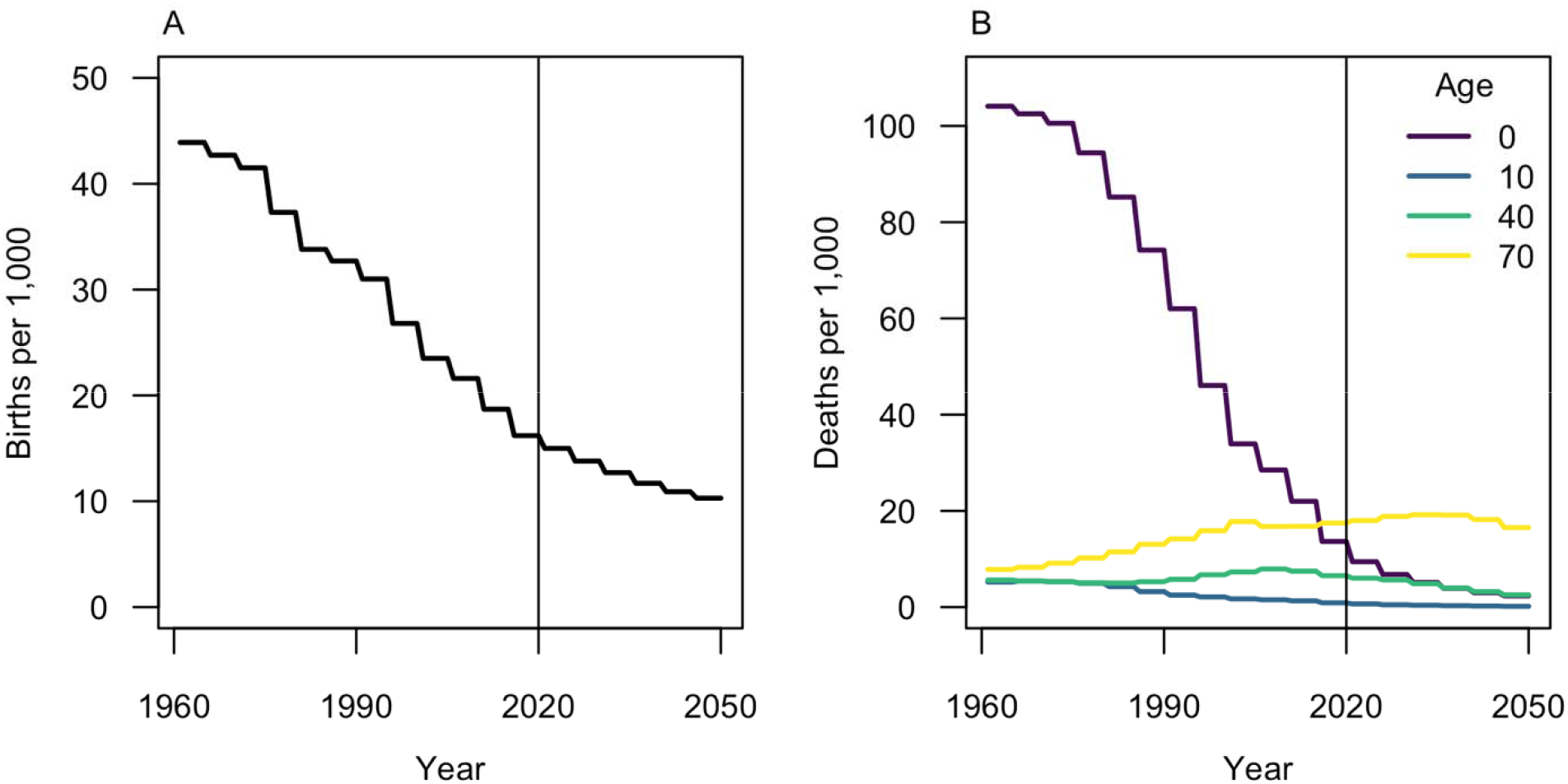
Demographic inputs over the period of model burn-in (left of vertical line) and post-vaccination (right of vertical line). A) Birth rate per 1,000 individuals. B) Death rate per 1,000 individuals, with four ages highlighted that span the diversity of patterns across ages.

#### DENV transmission

Like most models of mosquito-borne pathogen transmission (33), we did not explicitly incorporate mosquitoes into our model. Instead, the contribution of mosquitoes to DENV transmission was implicitly captured by transmission coefficients, *β*_*h*_ and *β*_*l*_. Within each community, we assumed that human-mosquito encounters were well-mixed and homogeneous, consistent with standard assumptions for ODE models. Between communities, we assumed that residents of one community spent a proportion, 1-*H*, of their time in the other community and were, therefore, subject to the force of DENV infection in that community during that time. Rather than model DENV serotypes explicitly, we assumed that the force of DENV infection was proportional to4-*s*, or the number of serotypes to which an individual had not yet been exposed. Individuals progressed through their period of active infection – and infectiousness to mosquitoes – at rate γ before progressing through a period of temporary cross-immunity at rate σ.

#### Dengue pathogenesis

Although the risk of severe disease is typically considered an important consideration for models of CYD-TDV vaccination impact, we limited our model to two types of infection: asymptomatic or symptomatic. Symptomatic and severe disease are often assumed to be highest among secondary infections, second highest among primary infections, and lowest among post-secondary infections (34). Hence, our model’s results about symptomatic disease may mirror what patterns of severe disease would be, had we modeled them.

To parameterize the proportion of infections that result in symptomatic disease as a function of *s, ρ*_*s*_, we adopted values used in a model by the Hopkins/UF team in Flasche et al. (2016) (Table 1). We also adopted their assumption that symptomatic infections are twice as infectious to mosquitoes as asymptomatic infections, despite substantial uncertainty around that multiplier (35). We chose to adopt values from this model due to its similarity to our model’s structure and its general consistency with seven other models of CYD-TDV vaccination impact (26).

#### CYD-TDV vaccination

In this analysis, we examined impacts of routine vaccination of nine-year-olds following a positive result from serological screening. For those in the *S*_*c,9,0*_ state, vaccination occurred at rate *v*_*c*_ (1 – *spec*). For those in the *S*_*c,9,1+*_, *I*_*c,9,1+*_, or *R*_*c,9,1+*_ states, vaccination occurred at rate *v*_*c*_ *sens*. Vaccinated individuals from the *S*_*c,9,s*_, *I*_*c,9,s*_, and *R*_*c,9,s*_ states then moved to state 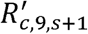. Then, they experienced a period of temporary cross-immunity to other DENV serotypes until they transitioned at rate σ to the appropriate 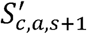 or *S*_*c,a,s+1*_ state.

Given our assumption that vaccination acts as a silent natural infection, there was no need for a parameter to describe protection afforded by vaccination, other than through the changes that result in rates of dengue pathogenesis as a consequence of *s* being incremented. For serological screening prior to vaccination, we assumed a test with sensitivity of 0.95 and specificity of 0.85 given the widely held view that both sensitivity and specificity would need to be high for CYD-TDV to have a positive impact (36). We held intervention coverage across the population as a whole at 50%, with that number reflecting a weighted average across two equally sized communities that each had intervention coverage ranging 0-100% across a range of scenarios that we considered. To explore the possible influence of a known trade-off between sensitivity and specificity in current DENV screening tests (37), we carried out additional analyses in which we varied specificity from 0 to 1 to identify the minimum value necessary to maintain positive benefits of the intervention.

#### Model equations

The aforementioned assumptions yielded the following equations that define the ODE model. Equations are presented in reference to community *c*, with the other community denoted as *c′*.

First, the dynamics of susceptible individuals of age *a* in community *c* followed

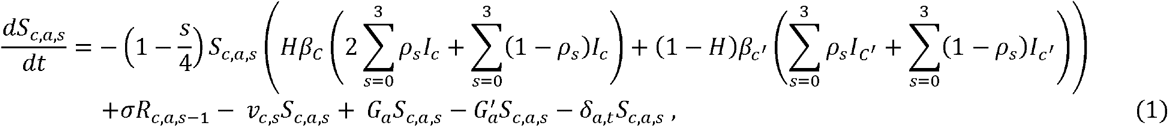

where the first term describes transmission from both communities weighted by the proportion of time *H* spent in each by a resident of *c*. Because individuals in this compartment have been exposed to *s* serotypes previously, the rate of infection of individuals in this class is reduced proportional to *s*. This formulation makes the simplifying assumption that there is equal circulation of the four DENV serotypes at any given time. Also within this first term of eqn. (1), contributions of infectious individuals are doubled for the proportion *ρ*_*s*_who are symptomatic. The second term in eqn. (1) indicates that vaccination occurs at rate v_*c,s*_, which is equal to (1 - *spec*) v_*c*_ for *s* = 0 and *sens* v_*c*_ for *s* > 0. In the third and fourth terms of eqn. (1), individuals age in (*G*_*a*_) and age out 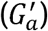 of each age class at a daily rate. In the fifth term of eqn. (1), deaths occur at the age- and time-specific rate δ_*a,t*_. For *s* = 0, µN_*c*_ is added to the susceptible state to represent births into the first susceptible class. In the sixth term of eqn. (1), individuals from the previous recovered class lose cross-protective immunity and enter the susceptible pool at rate σ, but only for *s* > 1.

Second, the dynamics of infected individuals of age *a* in community *c* followed

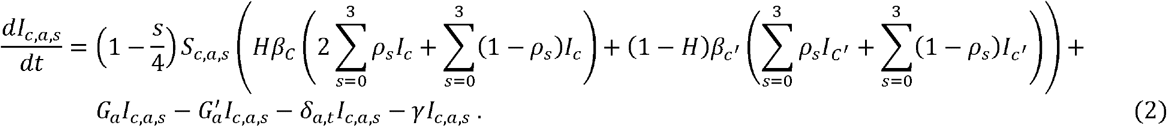

The first term in eqn. (2) is equal to the negative of the first term in eqn. (1), because they both describe the same transition. Similarly, the second, third, and fourth terms of eqn. (2) describe aging in and out of *I*_*c,a,s*_ and dying while being in that state. The fifth term of eqn. (2) describes individuals who recover from infection at rate γ.

Third, the dynamics of recovered individuals of age *a* in community *c* followed

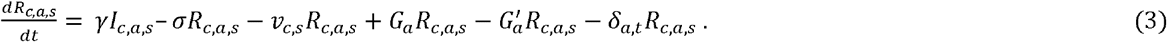

The only term in eqn. (3) that did not appear in eqns. (1) or (2) is the second term, which describes loss of cross-protective immunity at rate a.

Fourth, when individuals in S_*c,a,s*_ or R_*c,a,s*_ become vaccinated, they transition to state 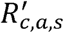, the dynamics of which follow

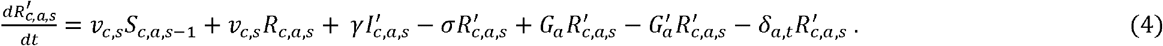

Eventually, vaccinated individuals lose cross-protective immunity at rate σ, entering state 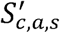, the dynamics of which follow

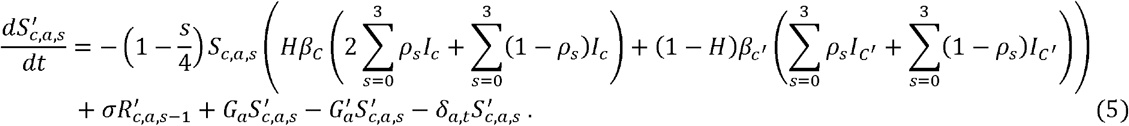

The dynamics of 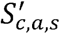 in eqn. (5) are similar to those of S_*c,a,s*_ in eqn. (1) except that there is no input to 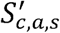 due to vaccination, and the force of infection has been reduced due to *s* effectively being incremented by 1 following vaccination. Individuals in state 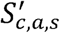 who become infected transition to state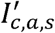, the dynamics of which follow

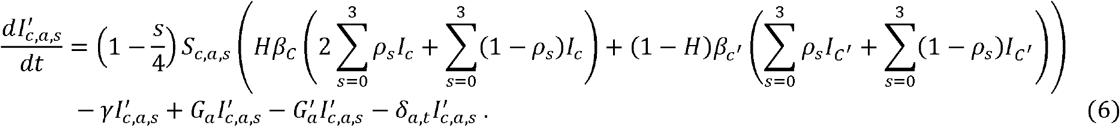

The dynamics of 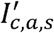 in eqn. (6) are effectively the same as those of *I*_*c,a,s*_ in eqn. (2).

#### Calibration

Following Flasche et. al (26), we used seroprevalence of nine-year-olds (SP9) as a proxy for DENV transmission intensity. Transmission coefficients were chosen to generate an SP9 of 20% in the low-transmission community and an SP9 of 80% in the high-transmission community. When both communities were evaluated together, this yielded an SP9 of 50%. This relatively wide range in SP9 was chosen purposefully, so as to explore the effects of a wide range in transmission intensities within a single, interconnected community. In particular, we chose a lower value of 20% so that a community in which CYD-TDV might possibly have a negative impact was considered.

To identify values of *β*_*h*_ and *β*_*l*_ that would yield the desired values of SP9 in the two communities, we solved the system of ODEs for sixty years under 100 different combinations of transmission coefficients for each community spanning a 10×10 grid of 0-0.2 for *β*_*l*_ and 0.2-0.3 for *β*_*h*_. To allow for interpolation between these values, we estimated the relationships between *β*_*h*_, *β*_*l*_, and SP9 using thin-plate splines. Values of *β*_*h*_ and *β*_*l*_ yielding values of SP9 of 20% and 80%, respectively, under mobility and no-mobility scenarios are provided in Table 1. Code for this calibration process is available at https://github.com/mwalte10/intra-urban_dengue_vaccination_impact/calibration.R.

### Analyses

We assessed the impacts of CYD-TDV vaccination by quantifying two metrics, proportion of cases averted and force of infection, described below. We did so separately in the two communities under scenarios with and without mobility and across varying levels of coverage allocated between the communities. We also performed the same calculations for a single, well-mixed community with SP9 = 0.5, which was intermediate between that of the low-(0.2) and high-transmission (0.8) communities.

#### Force of infection

To quantify the effects of the intervention on the overall dynamics of DENV transmission, we examined force of infection before and after introduction of CYD-TDV. We calculated force of infection as the rate at which susceptible people became infected on an annual basis, which corresponded to a weighted average of the first term in eqns. (2) and (6) across all *a* and *s* but stratified by *c* and *c’*.

#### Proportion of cases averted

Following previous work (13; 26), we used the proportion of symptomatic cases averted to quantify the impacts of CYD-TDV vaccination from a public health perspective. Negative values indicated an increase in cases, while positive values indicated a decrease. We applied this metric to vaccinated and unvaccinated populations separately to allow for the effects of direct and indirect protection to be assessed.

Because there is no corresponding group of vaccinated individuals in the model with no vaccination, we calculated the proportion of symptomatic cases averted by comparing equivalent fractions of age groups that were subject to vaccination in the model with vaccination.

## RESULTS

### Model validation

We compared the proportion of cases averted under our model to that under models used by Flasche et al. (26) at five baseline seroprevalence levels (10%, 30%, 50%, 70%, and 90%). As a function of baseline seroprevalence, the pattern of cases averted under our model was qualitatively similar to that under the other models (Fig. 3). Specifically, our model captured the same overall trend between baseline seroprevalence and cases averted and also predicted a switch from negative to positive cases averted as baseline seroprevalence increased from 10% to 30% (Fig. 3). Whereas this version of our model did not involve pre-vaccination screening (to be comparable to models used at the time by Flasche et al. (26)), the version of our model used to generate all subsequent results did involve pre-vaccination screening.

**Figure 3.**
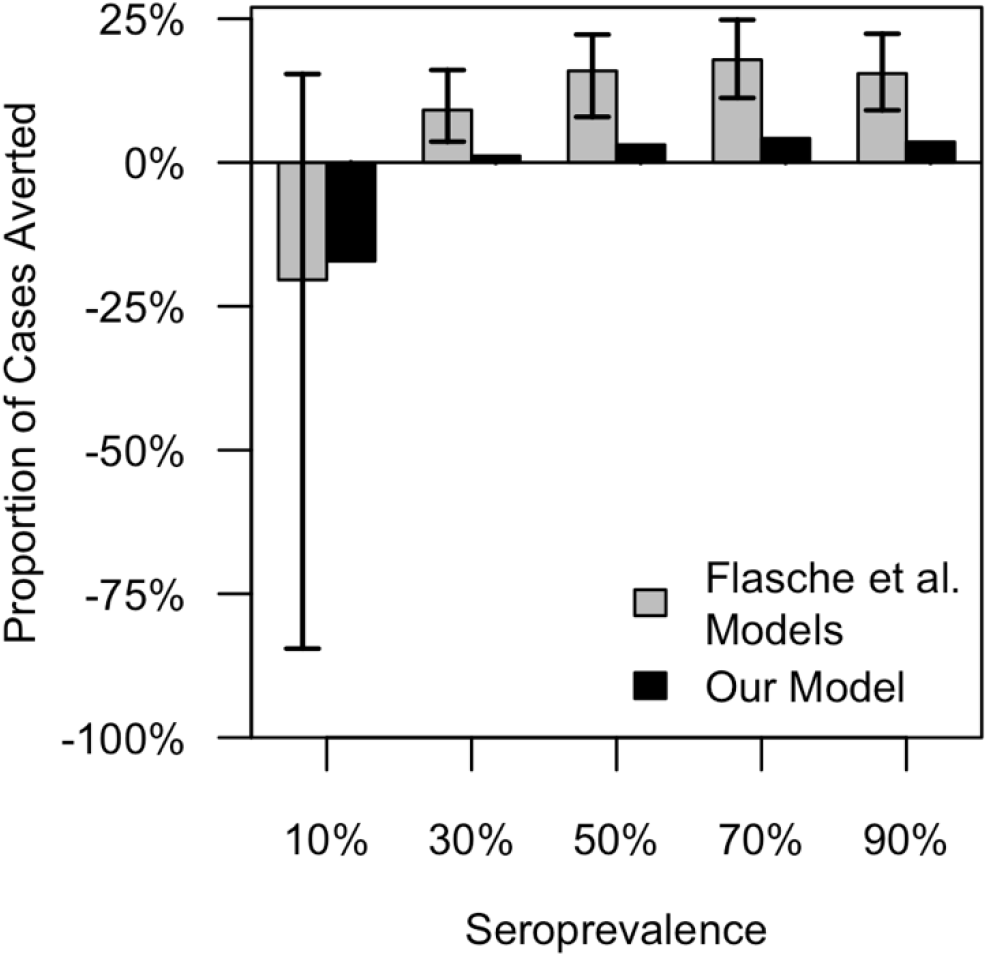
Model validation. We compared the proportion of cases averted predicted by our model to the range of predictions from eight models from Flasche et al. (26) across a range of values of baseline seroprevalence.

### Model behavior in the presence of vaccination

In all of our analyses, intervention coverage across the overall population was fixed at 50%, but the allocation of that coverage between the two communities varied. Vaccination coverage depended on intervention coverage, SP9 within a community, and the specificity and sensitivity of pre-vaccination screening. The highest vaccination coverage was achieved in the high-transmission community when 100% of the intervention was allocated there (Fig. 4). Under this scenario, 65.6% of the vaccination-eligible population in the high-transmission community was vaccinated. The low-transmission community generally had much lower vaccination coverage, with a peak of 14.9% coverage when 100% of the intervention was allocated there. In a version of the model with a single community with SP9 = 0.5, vaccination coverage was 21.2%, which was similar to vaccination coverage in the overall population when the intervention was allocated evenly across the two communities (Fig. 4).

**Figure 4.**
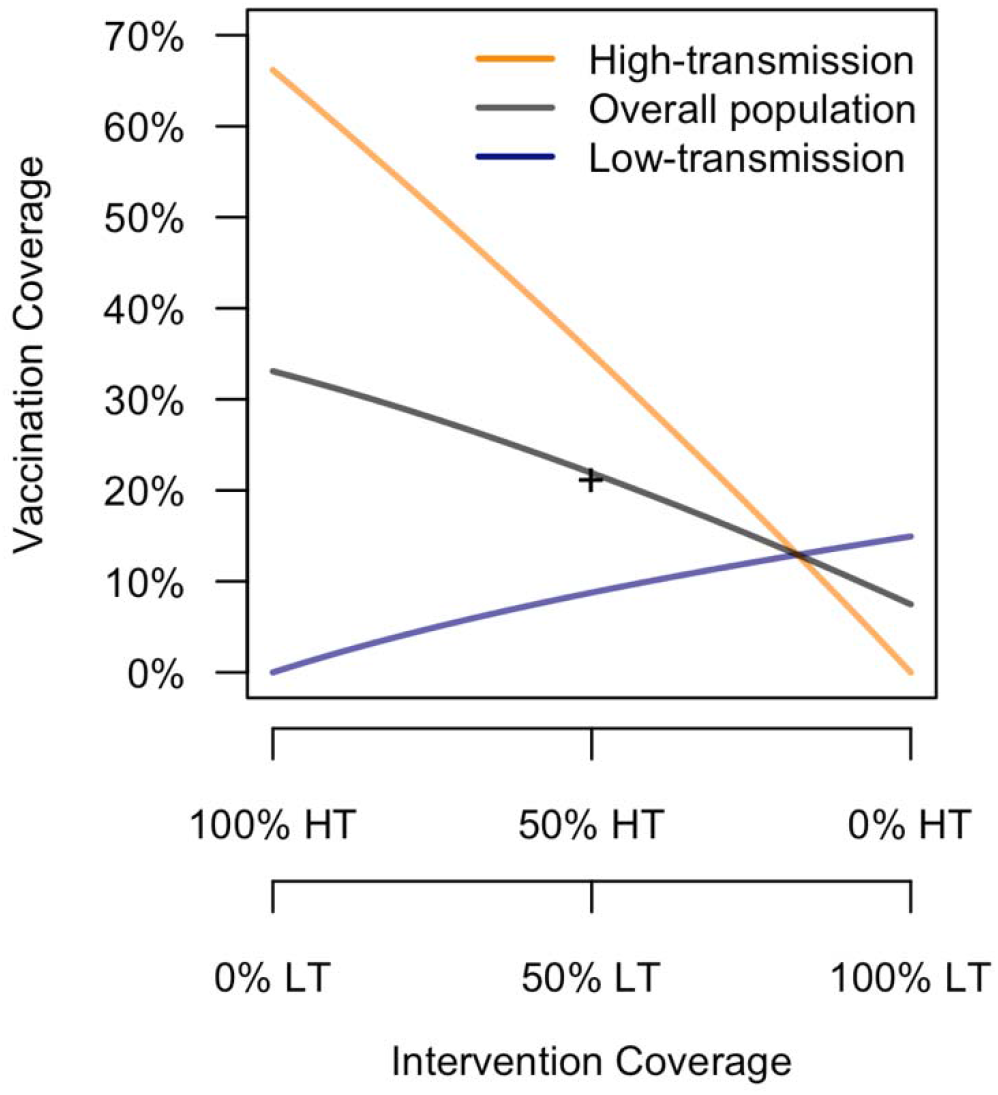
Dependence of vaccination coverage on intervention coverage. Vaccination coverage in each community (HT: high-transmission, LT: low-transmission) and the overall population (colored lines) was a function of intervention coverage in each community (x-axis), as well as the sensitivity and specificity of serological screening and seroprevalence among nine-year-olds, SP9 (fixed at default values). The + symbol indicates vaccination coverage in a single community with SP9 equal to the average of that of the low- and high-transmission communities.

Vaccination lowered the force of infection (FOI) in both communities, regardless of our assumptions about mobility (Fig. 5). When we assumed that there was no mobility between the two communities (Fig. 5A), each experienced its greatest decrease in FOI when it experienced higher intervention coverage. When we assumed that there was a modest degree of mobility between the two communities (Fig. 5B), both experienced greater decreases in FOI when intervention coverage was higher in the high-transmission community. This demonstrates that vaccination in the high-transmission community indirectly benefits the low-transmission community by reducing FOI there. Comparing the scenarios with and without mobility, the low-transmission community showed greater differences between these two scenarios than did the high-transmission community (Fig. 5), which was a result of transmission dynamics in the low-transmission community being primarily driven by the high-transmission community when there was mobility between the two communities. In a version of the model with a single community with SP9 = 0.5, the proportional reduction in force of infection was 6.0%, which was greater than in the high-transmission community but less than in the low-transmission community when the intervention was allocated evenly (Fig. 5).

**Figure 5.**
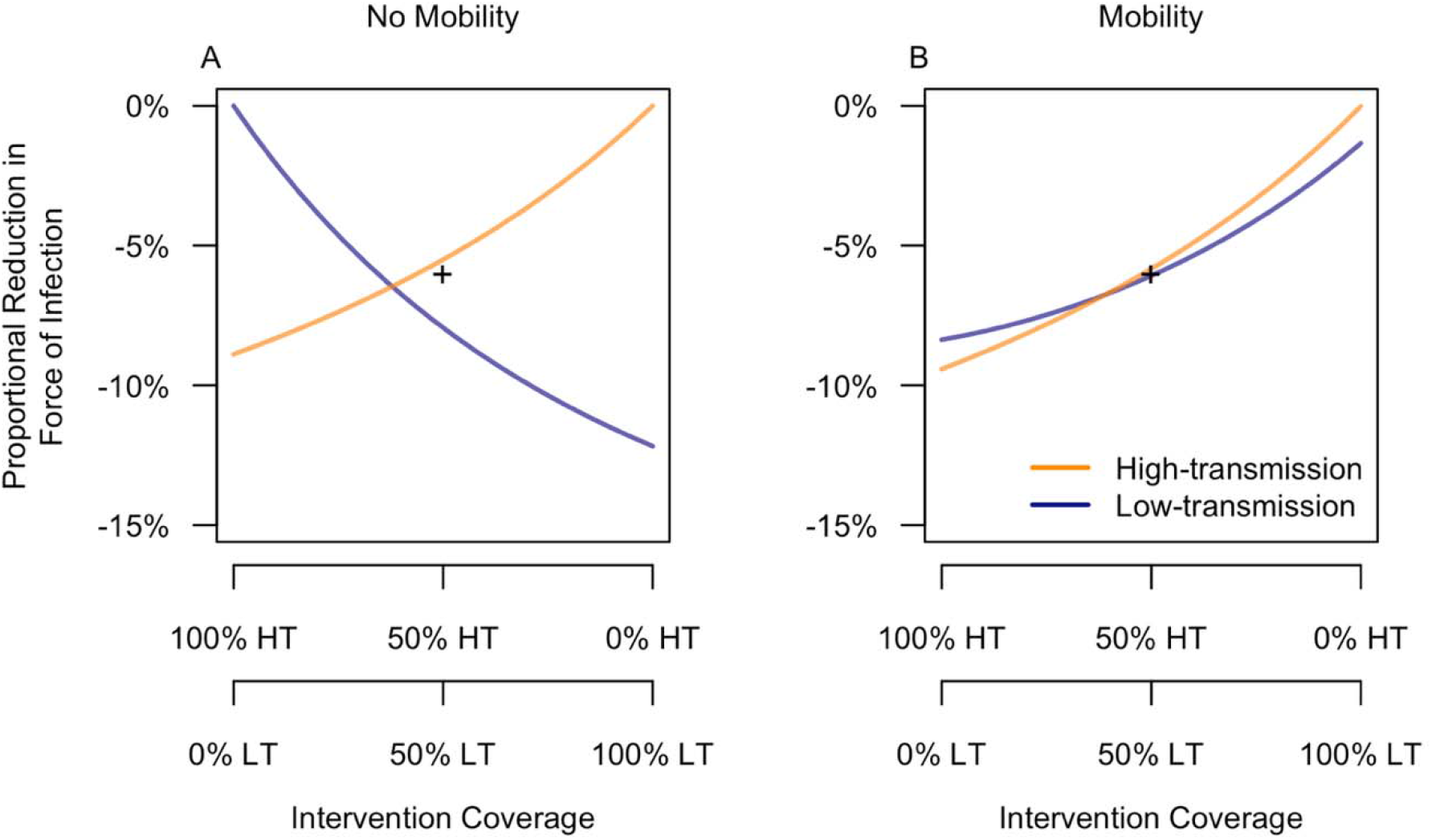
Proportional reduction in force of infection as a function of the allocation of intervention coverage across low- and high-transmission communities. Reductions are in reference to a baseline scenario with no intervention and are presented under scenarios in which the two communities (HT: high-transmission, LT: low-transmission) were or were not assumed to be connected through human mobility. The + symbol indicates the proportional reduction in force of infection in a single community with SP9 equal to the average of that of the low- and high-transmission communities.

### Vaccination impact in the absence of mobility

When the two communities were not connected through mobility, each experienced its greatest benefit from vaccination when it received 100% of the intervention (Fig. 6A). This mirrored reductions in FOI observed when each community received higher coverage (Fig. 5A). The population at large, however, experienced its greatest benefit when nearly 100% of the intervention was allocated to the high-transmission community (Fig. 6A). This is a result of the fact that the majority of infections occurred in that community. Because most of the population was unvaccinated, these trends for the overall population also applied to the unvaccinated population (Fig. 6C).

**Figure 6.**
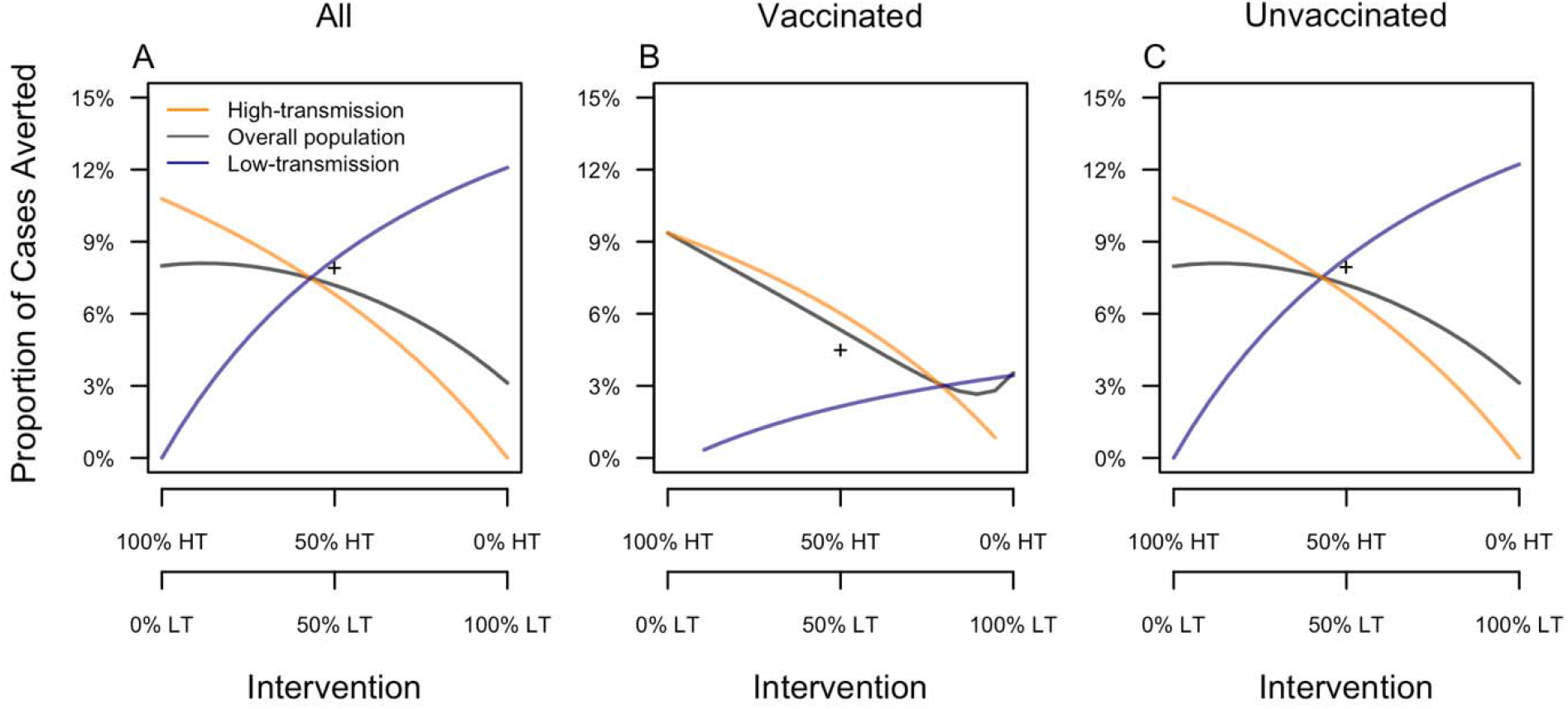
Proportion of cases averted under an assumption of no mobility between the two communities. Results are a presented as a function of the allocation of intervention coverage across the two communities (x-axis) (HT: high-transmission, LT: low-transmission) and broken down by vaccination status (columns) and community of residence (color). The + symbol indicates the proportion of cases averted in a single community with SP9 equal to the average of that of the low- and high-transmission communities.

Vaccinated individuals in a given community experienced their greatest benefit when 100% of the intervention was allocated to their home community (Fig. 6B). In the low-transmission community in particular, this was a consequence of our assumption of effective pre-vaccination screening, which ensured that the vaccine mostly went to those who stood to benefit from it (i.e., those for whom *s* > 0). The vaccinated individuals in the high-transmission community benefited more than those in the low-transmission community, because there were more individuals there for whom it was appropriate to receive CYD-TDV (Fig. 4). The two communities benefited equally when 79% of the intervention was allocated to the high-transmission community and 21% was allocated to the low-transmission community. Under different transmission parameters, we expect that this same pattern would hold but that the numerical values would differ.

One likely possibility for how the intervention might be deployed in practice is with even coverage across the two communities. Under this scenario, our model predicted that 6.6% of cases would be averted in the high-transmission community and 8.5% in the low-transmission community (Fig. 6A). Across the combination of the two communities, this amounted to an 11.7% reduction in the proportion of cases averted compared with the optimal scenario in which the intervention was allocated in a 79:21 ratio between the high- and low-transmission communities, respectively (Fig. 6A).

Among all individuals, the proportion of cases averted when the intervention was allocated evenly across the two communities was slightly lower than in a single community with SP9 = 0.5 (Fig. 6A). This was driven primarily by a higher proportion of cases averted among unvaccinated individuals in that community (Fig. 6C). In contrast, the single community with SP9 = 0.5 experienced a lower proportion of cases averted among vaccinated individuals (Fig. 6B).

### Vaccination impact in the presence of mobility

When the two communities were connected through mobility, the high-transmission community experienced its greatest benefit from vaccination when it received 100% of the intervention (Fig. 7A). The low-transmission community experienced its greatest benefit when it received 16% intervention coverage, at the expense of intervention coverage in the high-transmission community only being 84%. Compared to the scenario with 0% intervention coverage, increasing intervention coverage to 16% in the low-transmission community increased the proportion of cases averted there by only 0.2% (Fig. 7A). Because most of the population was unvaccinated, these trends for the overall population also applied to the unvaccinated population (Fig. 7C).

**Figure 7.**
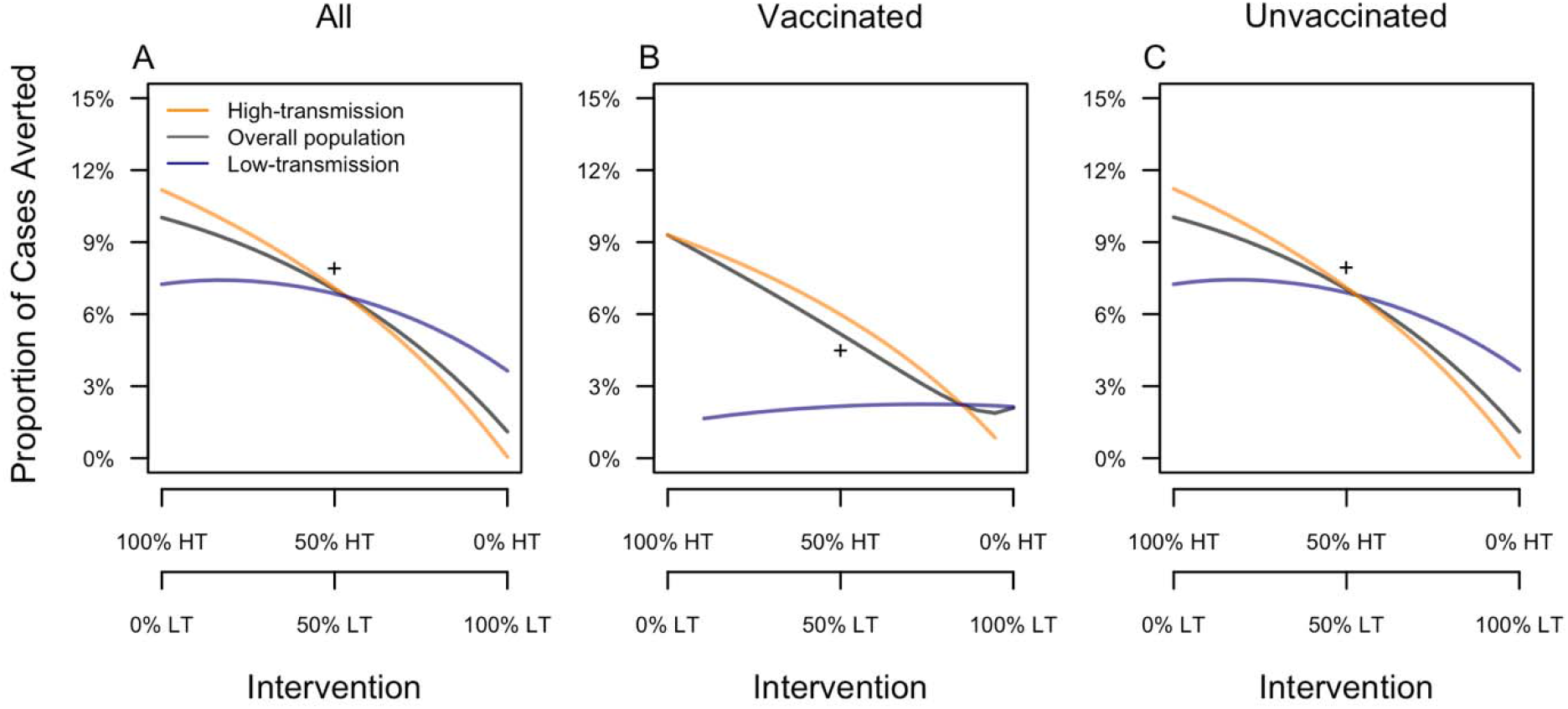
Proportion of cases averted under an assumption of mobility between the two communities. Results are a presented as a function of the allocation of intervention coverage across the two communities (x-axis) (HT: high-transmission, LT: low-transmission) and broken down by vaccination status (columns) and community of residence (color). The + symbol indicates the proportion of cases averted in a single community with SP9 equal to the average of that of the low- and high-transmission communities.

Vaccinated individuals in a given community experienced their greatest benefit when 100% of the intervention was allocated to their home community (Fig. 6E). This benefit amounted to 9.3% of cases averted in the high-transmission community and 2.6% of cases averted in the low-transmission community. Whereas reducing intervention coverage from 100% to 0% in the high-transmission community lowered the proportion of cases averted among vaccinated individuals there to nearly zero, reducing intervention coverage from 100% to 0% in the low-transmission community resulted in only a modest reduction in the proportion of cases averted among vaccinated individuals there (Fig. 7B).

Under the scenario of even intervention coverage across the two communities, there were considerably fewer cases averted in both the high- and low-transmission communities than under the scenario in which 100% of the intervention was allocated to the high-transmission community (Fig. 7A). Compared to the scenario with 100% of the intervention allocated to the low-transmission community, the scenario of even intervention coverage led to considerably more cases averted. Because most of the cases in the overall population derived from the high-transmission community, relative impacts of even and uneven intervention coverage scenarios for the overall population were similar to those for the high-transmission community (Fig. 7A).

Among all individuals, the proportion of cases averted when the intervention was allocated evenly across the two communities was slightly lower than in a single community with SP9 = 0.5 (Fig. 7A). This was driven primarily by a higher proportion of cases averted among unvaccinated individuals in that community (Fig. 7C). In contrast, the single community with SP9 = 0.5 experienced a lower proportion of cases averted among vaccinated individuals (Fig. 7B).

### Sensitivity analyses

Changes in the distribution of the overall population across the two communities resulted in modest, readily interpretable changes in our results about vaccination impact (Fig. 8). First, the proportion of cases averted in the high-transmission community was largely unaffected by whether more people resided there or in the low-transmission community (Fig. 8, purple lines across columns). Second, the proportion of cases averted in the low-transmission community was somewhat more sensitive to changes in the distribution of the population across the two communities (Fig. 8, yellow lines across columns), with the optimal intervention coverage in the low-transmission community being higher when a larger fraction of the overall population resided there. These first two observations can be explained by the fact that transmission in the low-transmission community was much more sensitive to the high-transmission community than the reverse. Third, the proportion of cases averted in the overall population more closely resembled the proportion of cases averted in whichever community had the larger population. This was primarily a consequence of how heavily the population-wide average was weighted by one community or the other.

**Figure 8.**
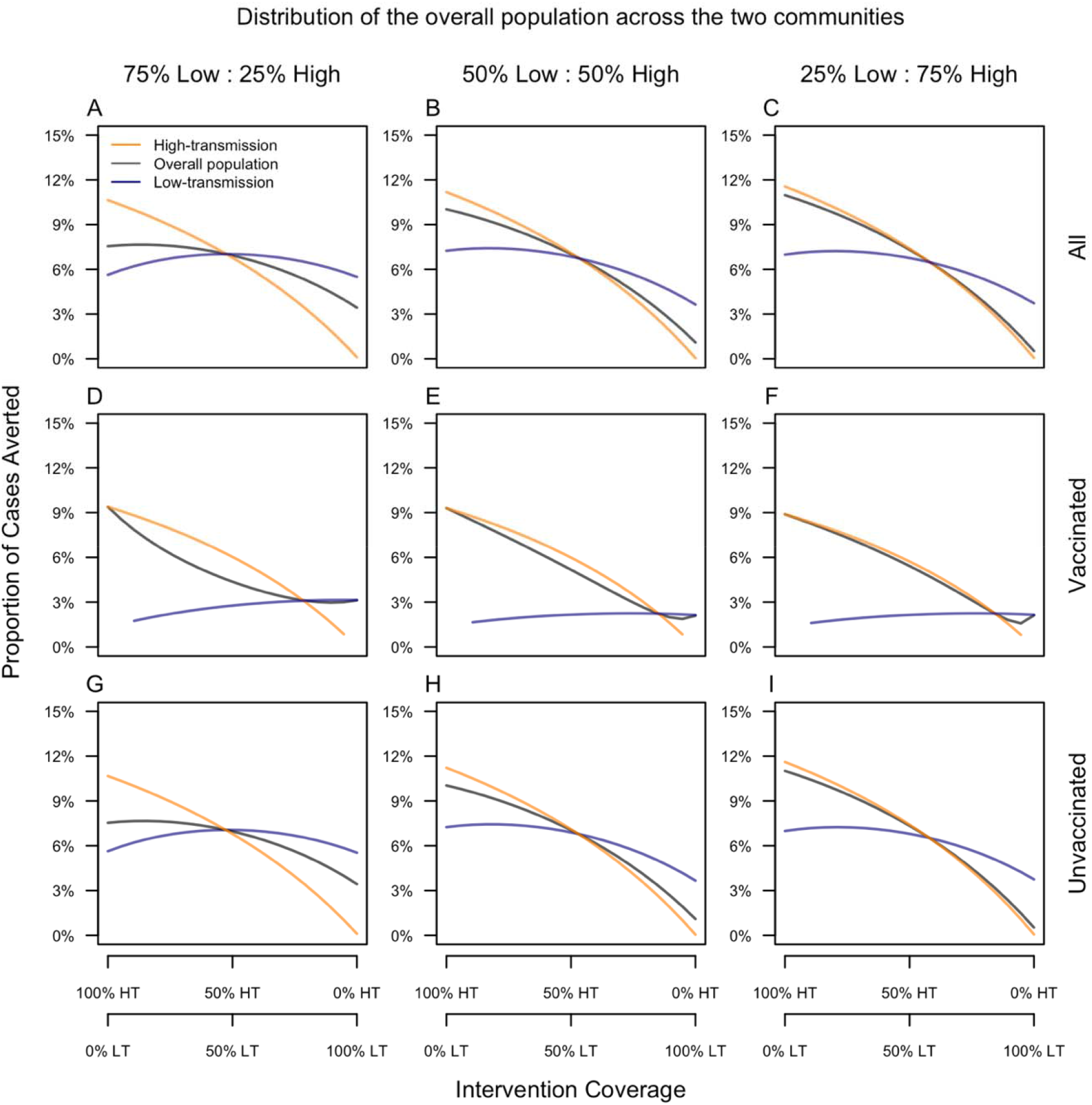
Sensitivity of the proportion of cases averted to unequal population sizes across the two communities (columns). Results are a presented as a function of the allocation of intervention coverage across the two communities (x-axis) (HT: high-transmission, LT: low-transmission) and broken down by vaccination status (rows) and community of residence (color).

Changes in the specificity of pre-vaccination screening had a substantial influence on our results about vaccination impact (Fig. 9). Sensitivity of the proportion of cases averted to screening specificity was greatest among vaccinated individuals, with extremely low values of specificity resulting in a negative proportion of cases averted in both low- and high-transmission communities (Fig. 9C & 9D). These effects did differ somewhat across the two communities though, with the decline in the proportion of cases averted associated with lower values of specificity being much steeper in the low-transmission community (compare spacing between lines in Fig. 9C & 9D). The proportion of cases averted among unvaccinated individuals was also sensitive to screening specificity, but mostly in the low-transmission community (Fig. 9C). This was a consequence of there being lower seropositivity among potential vaccine recipients in the low-transmission community, which meant that lower values of screening specificity had a greater impact on vaccination coverage there than in the high-transmission community.

**Figure 9.**
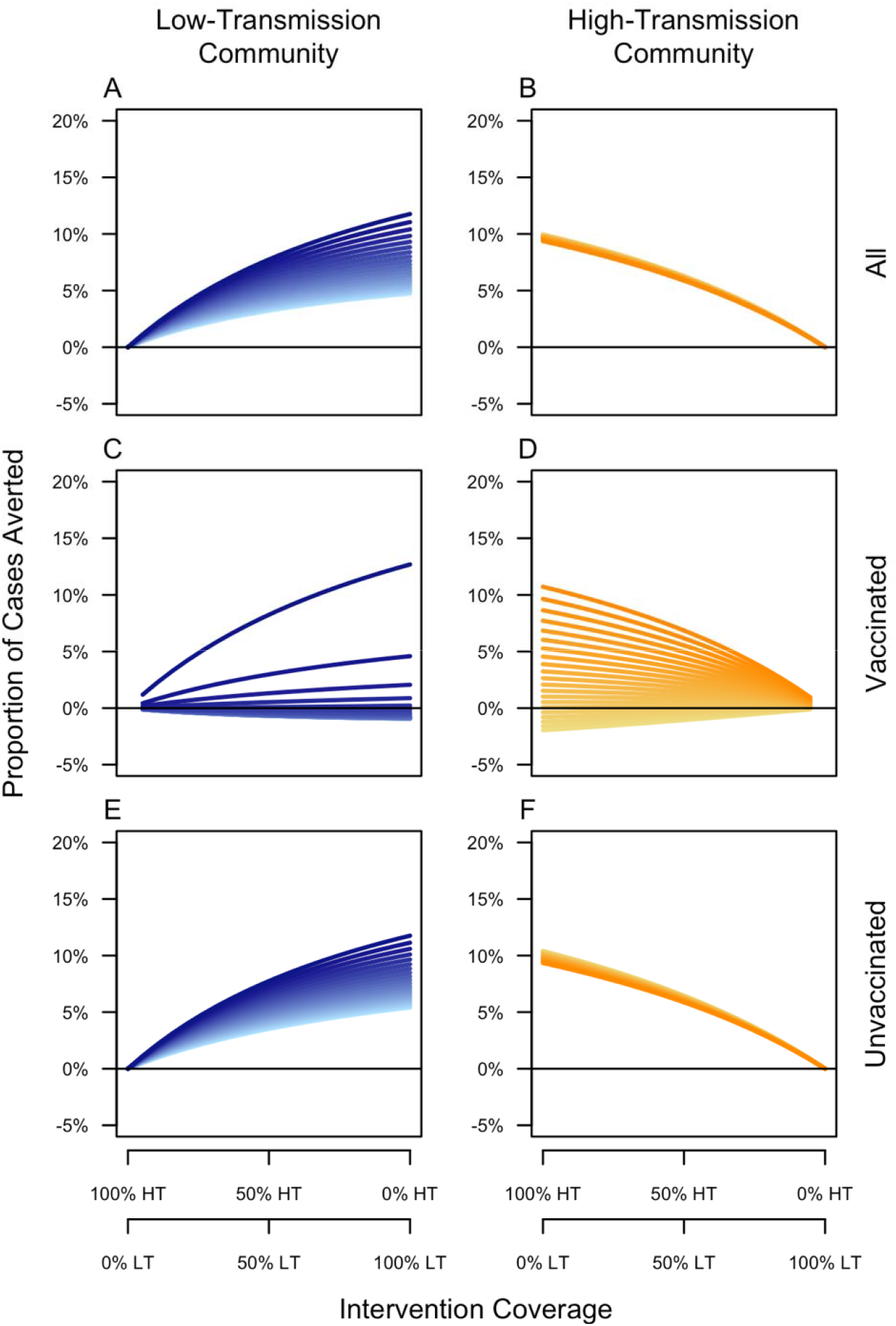
Sensitivity of the proportion of cases averted to the specificity of pre-vaccination screening. Results are a presented as a function of the allocation of intervention coverage across the two communities (x-axis) (HT: high-transmission, LT: low-transmission) and broken down by vaccination status (rows) and community of residence (columns). Specificity values range 0-1, with lighter colors representing lower values and darker colors representing higher values.

## DISCUSSION

Although other models that feature spatial transmission heterogeneity (38; 39; 40; 41) have been used to project CYD-TDV vaccination impact (26), our study is the first to directly assess the influence of spatial transmission heterogeneity on projections of CYD-TDV vaccination impact. To ensure that our model’s behavior was comparable to that of other models, we compared its predictions about vaccination impact to predictions from eight other models (26) that influenced the WHO’s initial policy recommendation about this vaccine (8), finding comparability between our model and these others in key respects. Our model’s behavior was also sensible in that vaccination coverage in each community depended on the seropositivity of vaccinees in each (11), and reductions in force of infection mirrored increases in vaccination coverage. Based on these results, we deemed our model suitable for making an initial, theoretical assessment of the impacts of CYD-TDV vaccination in a spatially heterogeneous population.

In the event that two or more communities are relatively isolated from one another (e.g., cities within a state), our results based on a model with no mobility indicate that each community would benefit maximally if 100% of the intervention were targeted to itself. From the perspective of the overall population (i.e., the state as a whole rather than each city), targeting the intervention to the community with the highest transmission would have the greatest impact. In the event that two or more communities experience an appreciable degree of coupling (e.g., neighborhoods within a city), our results based on a model with mobility indicate that targeting the intervention to the community with the highest transmission would have the greatest impact, both from the perspective of the overall population and from the perspective of each community. Given that spatial coupling has been found to be important for DENV transmission at multiple spatial scales (42; 43; 44; 45), taken together, our results suggest that spatial targeting could enhance the impact of vaccination with CYD-TDV following pre-vaccination screening. Doing so successfully would require overcoming a number of challenges to implementation, including more spatially detailed estimates of DENV seropositivity (22; 46) and more accurate assays for serological screening (47).

Because spatial targeting might not always be feasible, we also considered a scenario involving even intervention coverage across the two communities. Based on a model with no mobility, intervention impact under even coverage was only slightly suboptimal from the perspective of the overall population. Based on a model with mobility, however, intervention impact under even coverage was considerably less than under the optimal scenario of targeting the intervention exclusively on the high-transmission community. Moreover, from the perspective of vaccinated individuals, even intervention coverage across the two communities was always suboptimal, as it reduced vaccination coverage among seropositive individuals in the high-transmission community who would have benefited from vaccination (9). At the same time, impact under even intervention coverage was considerably higher than under scenarios in which the intervention was allocated preferentially to the low-transmission community. Due to the possibility that socioeconomic factors could underlie a coupling between heterogeneities (48) in DENV transmission (19; 20; 21; 22) and access to dengue vaccines (24; 25; 49), it will be important for measures to be taken to ensure that coverage of CYD-TDV following pre-vaccination screening is no less than even, if not targeted preferentially to communities that contribute disproportionately to transmission.

Projections of CYD-TDV vaccination impact to date have mostly assumed even coverage in a population in which transmission is spatially homogeneous (12). Agent-based models have been an exception in that they have usually accounted for spatial heterogeneity to some extent, although they too have neglected the issue of uneven coverage and have incorporated spatial heterogeneity in ways that are complicated and highly specific to a given location (38; 39; 40; 41). Comparison of versions of our model with and without spatial heterogeneity offers simple and straightforward intuition about whether ignoring spatial heterogeneity might lead to bias in CYD-TDV impact projections. Overall, our results indicate that models based on a single, spatially homogeneous population may produce reasonable projections of impact in a spatially heterogeneous population under even intervention coverage. At the same time, we did observe tendencies for the model based on a single population to project higher impact in unvaccinated individuals and lower impact in vaccinated individuals than did the model based on spatially heterogeneous communities. Most significantly though, ignoring spatial heterogeneity led to substantial overestimation of impact in vaccinated individuals in the low-transmission community, which is problematic given safety concerns about CYD-TDV in seronegative individuals (9; 50). Combined with indications from our sensitivity analysis that low screening specificity could lead to an increase in cases among vaccinated individuals in the low-transmission community, this suggests that there may be scenarios in which models assuming spatial homogeneity predict positive impacts that are, in reality, negative for a segment of the population.

Although the relatively simple model that we used here is conducive to the development of intuition about the influence of spatial heterogeneity on CYD-TDV vaccination impact, it leaves open a number of questions about this issue under more realistic circumstances. To begin to address these questions, we performed two sets of sensitivity analyses. The first indicated that a model with uneven population distribution across high- and low-transmission communities would behave similar to our model with even population distribution except that the weighting of impact across the two communities would follow the distribution of population across the two communities. The second indicated that impact in the low-transmission community was more sensitive to the specificity of serological screening than was impact in the high-transmission community, although the allocation of intervention coverage across the two communities also affected the sensitivity of vaccination impact to the specificity of serological screening. Even so, the generality of our results would best be assessed through understanding of the extent to which our conclusions hold when analyzed in a similar fashion by other models in more realistic settings. Similar follow-up analyses have occurred from the perspective of estimating CYD-TDV cost-effectiveness (51; 52; 53). One way that this could be achieved in practice is if future studies involving models of CYD-TDV vaccination impact were to perform sensitivity analyses on the issue of spatial heterogeneity in DENV transmission.

In practice, the spatial scale at which decisions about CYD-TDV vaccination will be made may be as coarse as a country or the first administrative level within a country (e.g., state, province) (46). Given that DENV transmission is typified by spatial heterogeneity below those scales (15; 16; 17; 18; 22), our results are pertinent to future modeling projections of the impact of CYD-TDV vaccination policies at those scales. Although we have shown that projections ignoring spatial heterogeneity may provide reasonably accurate insights about overall impact under even intervention coverage, there are other, potentially serious, limitations of those approaches. First, in the event that there is capacity to target the intervention to high-transmission communities, models that ignore spatial heterogeneity will underestimate impact. Second, in the event that socioeconomic drivers or other factors result in unintentional targeting of low-transmission communities, models that ignore spatial heterogeneity will overestimate impact. Third, even in the event of even intervention coverage, models that ignore spatial heterogeneity could substantially overestimate impact, or even overlook the potential for harm, among vaccinated individuals in low-transmission communities. Further developments in modeling and additional collection and sharing of fine-scale spatial data will be important for addressing these issues and maximizing the potential benefits of this dengue vaccine, and potentially others that come after it (36; 54).

## Data Availability

All code and data is available at https://github.com/mwalte10/intra-urban_dengue_vaccination_impact

## ACKNOWLEDGEMENTS

We thank members of the Perkins Lab for feedback on this work.

